# Caregiver knowledge and Attitudes toward Human Papillomavirus Vaccination among adolescent girls living with HIV in Sierra Leone: A health facility-based cross-sectional study

**DOI:** 10.64898/2026.09.21.26363609

**Authors:** Darlinda F Jiba, Mamadu Baldeh, Patrick Turay, Saidu Kanu, Matilda N Kamara, Lynda M. L Farma, Umu Barrie, Waheed O Awonuga, Mary M Baio, Daniel Sesay, Diana Shehab, Rosaline Sinnah, Enanga S Namanga, Phildys M D Johnson, Sulaiman Lakoh, J. Andrew Dykens

## Abstract

**Objective:** To assess the knowledge, willingness and barriers to human papillomavirus (HPV) vaccination among caregivers of adolescent girls living with HIV in Sierra Leone.

**Methods:** This cross-sectional study examined HPV vaccine knowledge, willingness, and barriers among 249 caregivers of adolescent girls living with HIV in Sierra Leone. Data were collected through structured questionnaires at public tertiary hospitals and high-volume HIV clinics, and associations were assessed using chi-square tests, logistic regression, and Kruskal-Wallis analyses.

**Results:** Only 32.1% (80/249) of caregivers had heard of HPV, with even fewer aware of its link to cervical cancer (24.4%, 60/246) or prevention methods of cervical cancer (31.9%, 79/248). Misconceptions were prevalent; 16.3% (36/221) erroneously linked oral contraceptives to cervical cancer, and 61.1% (44/72) of caregivers unwilling to vaccinate cited distrust in vaccine efficacy. While tertiary-educated caregivers exhibited the highest level of knowledge (45.5%, p < 0.001), their willingness to vaccinate (50%, 33/66) was comparable to that of less-educated groups, highlighting a knowledge-action gap. Structural barriers dominated, with 66.1% (154/233) citing unawareness of the vaccine and 10.3% (24/233) missing school-based vaccination days. Relationship dynamics significantly influenced willingness: extended family members showed the highest willingness (69.4%, p = 0.029), whereas grandparents were least willing (31%, 9/29). Although systemic gaps such as disrupted vaccination services, vaccine supplies and cold chain limited uptake, healthcare workers emerged as the most effective information source (rank sum = 889.5, p = 0.039).

**Conclusion:** The study highlights the need for culturally tailored education to dispel myths and community-driven strategies engaging trusted actors like healthcare workers and extended families. This study justifies the need for integration of HPV vaccination and cervical cancer screening into HIV care services.

## Background

Cervical cancer, a malignancy affecting the cervix, is the fourth most common cancer and cause of cancer-related mortality among women worldwide [1, 2]. This burden disproportionately affects low-resource regions, where approximately 90% of cases occur, with mortality rates doubling those observed in high-income countries [2,3,4]. Sub-Saharan Africa (SSA), in particular, bears a high burden, accounting for 70% of global cervical cancer cases [4, 5].

Human papillomavirus (HPV), a sexually transmitted infection, is the primary causative pathogen of cervical cancer [6, 7]. The global prevalence of HPV is highest in SSA, with an estimated 24% of women infected, particularly unvaccinated, sexually active adolescents and young women under 25 years [8,9,10]. Compounding this risk is the synergistic relationship between HPV and human immunodeficiency virus (HIV). HIV-induced immunosuppression, marked by low CD4+ T-cell counts and elevated viral loads, heightens susceptibility to persistent HPV infection and accelerates progression to malignancy [11, 12]. Studies confirm HIV as a significant risk factor for HPV persistence and cervical carcinogenesis, underscoring the vulnerability of HIV-seropositive populations [13, 14].

The World Health Organization (WHO) advocates a multi-strategic approach to cervical cancer prevention including mass HPV vaccination, HPV vaccination for girls before sexual debut and screening and treatment of precancerous lesions in women[15]. Despite the vaccine’s proven safety and efficacy in mitigating HPV persistence, global coverage remains inequitable. Only 40% of women worldwide are vaccinated, with 70% of doses administered in high-income countries compared to 2.7% in low- and middle-income countries (LMICs) [16, 17]. Beyond access barriers, vaccine hesitancy, driven by misinformation and caregiver decision-making, further impedes uptake. As primary decision-makers for minors, caregivers critically influence vaccination success, acceptance in this age group hinges on disease awareness, information accessibility, and perceived vaccine safety.

In Sierra Leone, HPV vaccination targets girls aged ≤10 years through school-based programs. However, there is a gap in the evidence on caregiver knowledge and willingness to vaccinate adolescent girls living with HIV (AGLHIV) against HPV. The purpose of this article is to report the findings from a nationwide study that engaged caregivers on knowledge, attitudes, and willingness regarding HPV vaccination for adolescent girls living with HIV in Sierra Leone, aiming to inform strategies for integrating cervical cancer screening in HIV care programs.

## Methods

### Study design and study setting

We conducted a national cross-sectional study involving caregivers of adolescent girls living with HIV attending public hospitals and high-volume antiretroviral therapy (ART) clinics (ART clinics with more than 1,000 clients) providing specialized adolescent and child HIV services in Sierra Leone

Sierra Leone’s healthcare system is structured across three tiers: primary, secondary, and tertiary care. Under the National AIDS Control Programme, 675 ART clinics operate nationally, with tertiary care delivered at regional and national hospitals. Facility selection included all three regional hospitals and five additional high-volume ART clinics offering paediatric and adolescent HIV care.

### Study population, sample size and sampling methods

This study enrolled caregivers of AGLHIV aged 10–19 years receiving care at public tertiary hospitals or high-volume ART clinics in Sierra Leone. Caregivers were defined as parents, legal guardians, or primary adults responsible for the adolescents’ health decisions. Participants were recruited between August 1, 2023, and June 1, 2024, during routine clinic visits. Inclusion criteria required caregivers to be ≥18 years old, residing with the AGLHIV for ≥6 months, and providing informed consent. Individuals with cognitive impairments or language barriers precluding informed consent were excluded.

The designated ART clinic nurses generated a list of all AGLHIV and provided contact details for their caregivers. Identified caregivers who met the inclusion criteria were contacted and invited to participate in the study. Sample size estimation was conducted using Epi-Info™ version 7.2.5 (CDC, Atlanta, GA), with parameters derived from prior regional studies. For the primary outcome (caregiver knowledge of HPV vaccination), we assumed a baseline knowledge prevalence of 58.4%, while the secondary outcome (willingness to vaccinate) was based on an anticipated prevalence of 42%. Adjusting for a 10% non-response rate, the minimum sample sizes were 150 (knowledge) and 100 (willingness). The larger estimate (n = 210) was selected to ensure statistical power for both outcomes.

### Data Collection Procedures

A structured questionnaire, administered via face-to-face engagement, was employed to collect data. The instrument was developed in English and executed in Krio (the local lingua franca). Data collectors were recruited from certified HIV nursing staff proficient in Krio and experienced in adolescent HIV care protocols. Collectors were trained in informed consent, confidentiality, and non-coercive communication. The study investigator re-interviewed 10% of participants to verify response consistency (κ > 0.85). A pretest involving 10 caregivers from a non-participating clinic informed refinements to question phrasing and response options. Post-pretest revisions enhanced internal consistency, increasing Cronbach’s α from 0.72 to 0.81 for knowledge items.

### Data management and analysis

The data were collected using paper-based questionnaires, entered into Epi-Data version 4.2. All participant identifiers were replaced with anonymized codes to protect confidentiality. Following daily data collection, encrypted backups were stored on password-protected servers accessible only to the principal investigators. Missing data were analyzed for patterns using Little’s MCAR test. Variables with <5% missingness were addressed via complete case analysis, while those exceeding this threshold were imputed using multivariate chained equations (MICE).

Analyses were conducted in Stata SE 18.0 (StataCorp, College Station, TX), with statistical significance set at p < 0.05 (two-tailed). Categorical variables (e.g., caregiver sex, residential setting) were summarized as frequencies and percentages and continuous variables (e.g., knowledge scores) were assessed for normality using Shapiro-Wilk tests and reported as means (standard deviations) or medians (interquartile ranges). Chi-square or Fisher’s exact tests compared proportions of adequate knowledge (≥70% correct responses) and willingness (Likert score ≥4) across sociodemographic strata. Kruskal-Wallis tests evaluated differences in knowledge scores by information sources.

Two hierarchical logistic regression models were constructed to examine associations between adequate knowledge (dependent variable) and sociodemographic factors, information sources, and health literacy and to assess predictors of willingness to vaccinate, adjusting for knowledge, sociodemographics, and prior vaccination history (Table 1). This model building followed Hosmer-Lemeshow’s stepwise approach, where unadjusted associations (p < 0.25) were retained, entered into the model using backward elimination (p < 0.05) and adjusted odds ratios (aORs) with 95% confidence intervals (CIs) were computed. Multicollinearity was assessed via variance inflation factors (VIF < 5).

**Table 1:** Study variables and operational definitions.

| **Dependent Variables:** |  |
| --- | --- |
| Caregiver Knowledge of HPV Vaccination | Operationalized as the proportion of correct responses to 12 validated items assessing understanding of HPV transmission, cervical cancer etiology, and vaccine efficacy. |
| Willingness to Facilitate HPV Vaccination | Measured on a 5–point Likert scale (1 = strongly unwilling to 5 = strongly willing), with willingness defined as a score $\geq 4$. |
| Independent Variables |  |
| Sociodemographic Covariates | Caregiver age (categorized as 18–30, 31–45, >45 years)<br>Sex (male/female)<br>Relationship to the adolescent (biological parent, relative, non-kin guardian)<br>Educational attainment (none, primary, secondary, tertiary)<br>Occupation (employed/unemployed)<br>Residential setting (urban/rural)<br>Religious affiliation<br>Monthly household income (<50, 50–100, >100,>100)<br>Prior childhood vaccination history (yes/no) |
| Information Exposure Pathways | Sources of health information:<br>• Mass media (radio, television, newspapers)<br>• Digital platforms (social media)<br>• Interpersonal networks (healthcare providers, religious leaders, family members, community peers). |
|  | Exposure was dichotomized (exposed/unexposed) based on self-reported access. |
| Health Literacy and Perceptions | Understanding of HPV infection risks, cervical cancer prevention, and vaccine safety/benefits, assessed via 10-item scales adapted from WHO's Vaccine Hesitancy Survey Toolkit [18, 19]. |
| Operational Definitions |  |
| Awareness | Defined as caregivers' self-reported exposure to information related to HPV, cervical cancer, or HPV vaccination. Awareness was operationalized as attentiveness to and recognition of these topics, irrespective of factual accuracy. |
| Knowledge | Assessed as the proportion of factually correct information caregivers retain about HPV transmission, cervical cancer etiology, and vaccine efficacy. |

### Ethical Approval and Participant Consent

Ethical clearance for this study was obtained from the Sierra Leone Ethics and Scientific Review Committee (SLESRC Approval No.: 022/07/2023). Administrative authorization was secured from participating in antiretroviral therapy (ART) clinics through formal institutional agreements, permitting access to de-identified programmatic data collected during routine service delivery. Before enrollment, written informed consent was obtained following a structured disclosure process. Trained staff verbally explained the study’s objectives, risks, benefits, and participants’ rights in Krio, the local lingua franca. Consent documentation emphasized the voluntary nature of participation and the right to withdraw at any time without affecting clinical care.

To ensure confidentiality, all participant identifiers were replaced with pseudonymized codes during data entry. Aggregated reporting and secure storage protocols (password-protected, encrypted servers) were implemented to prevent re-identification. Clinic staff were not involved in recruitment or data collection to mitigate coercion risks.

## Results

### Baseline sociodemographic characteristics

Table 2 presents findings of the sociodemographic characteristics of 249 caregivers of AGLHIV attending 8 high volume ART clinics in Sierra Leone stratified by HPV vaccination status. A mean duration of HIV diagnosis (17.79 months, SD = 15.47) was noted among vaccinated AGLHIV compared to unvaccinated (42.9 months, SD = 48.74; p = 0.008). Females constituted the majority (78.6%, 195/249) of caregivers, with comparable vaccination rates (21.5%, 42/195) to male caregivers (17.6%, 9/51). Vaccinated adolescents were slightly older (median = 16 years, IQR = 15–18) than unvaccinated adolescents (median = 14 years, IQR = 11–16). Caregivers who were extended family members account for the highest vaccination rate among ALHIV (25%, 9/36), followed by family friends (18.2%, 2/11). In contrast, grandparents (0%, 0/29) and siblings (7.3%, 3/41) had the lowest rates (p = 0.023) of vaccination rates. Employed caregivers demonstrated higher vaccination rates (15.6%, 31/199) compared to unemployed caregivers (4.8%, 2/42; p < 0.010).

**Table 2.**
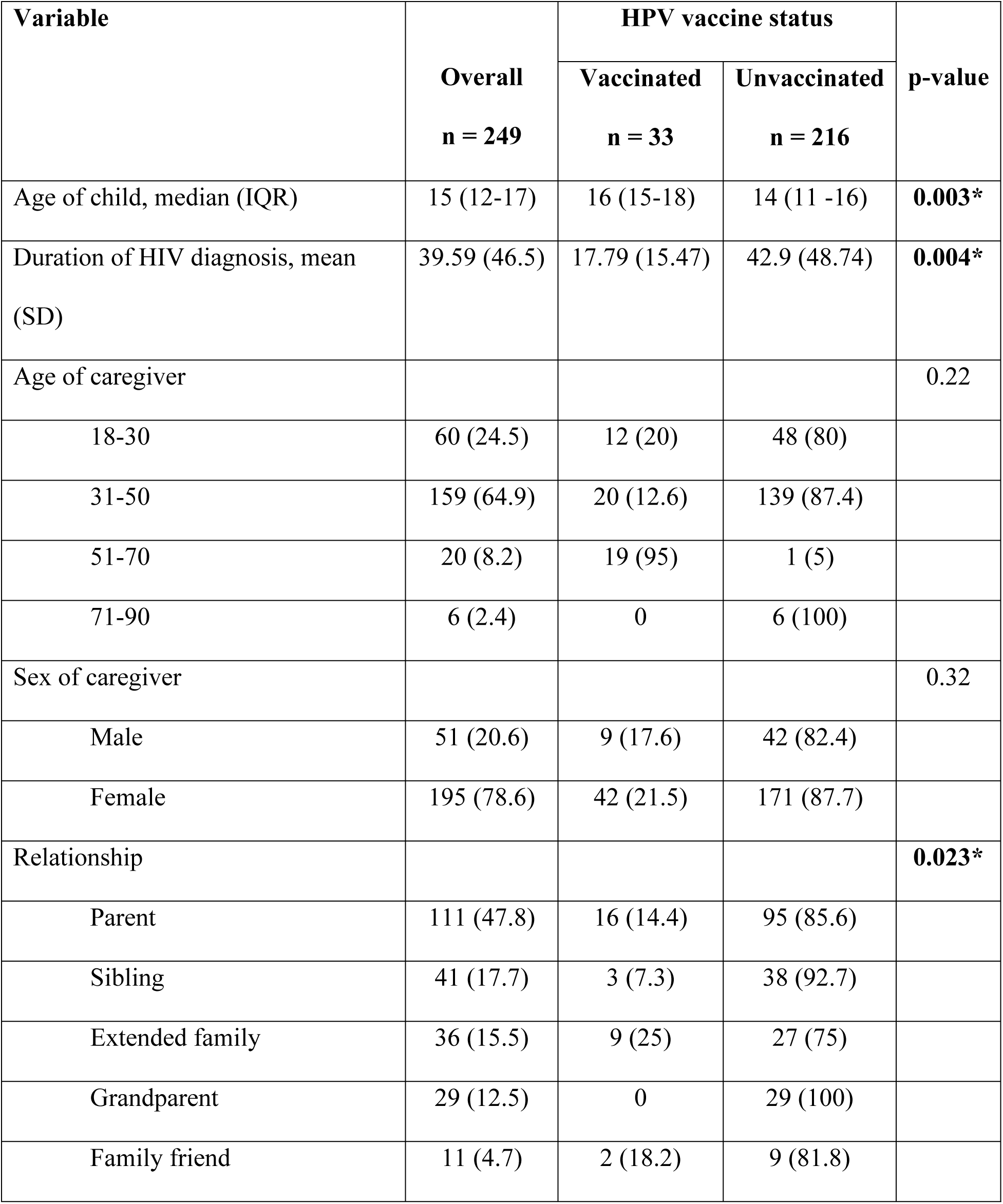
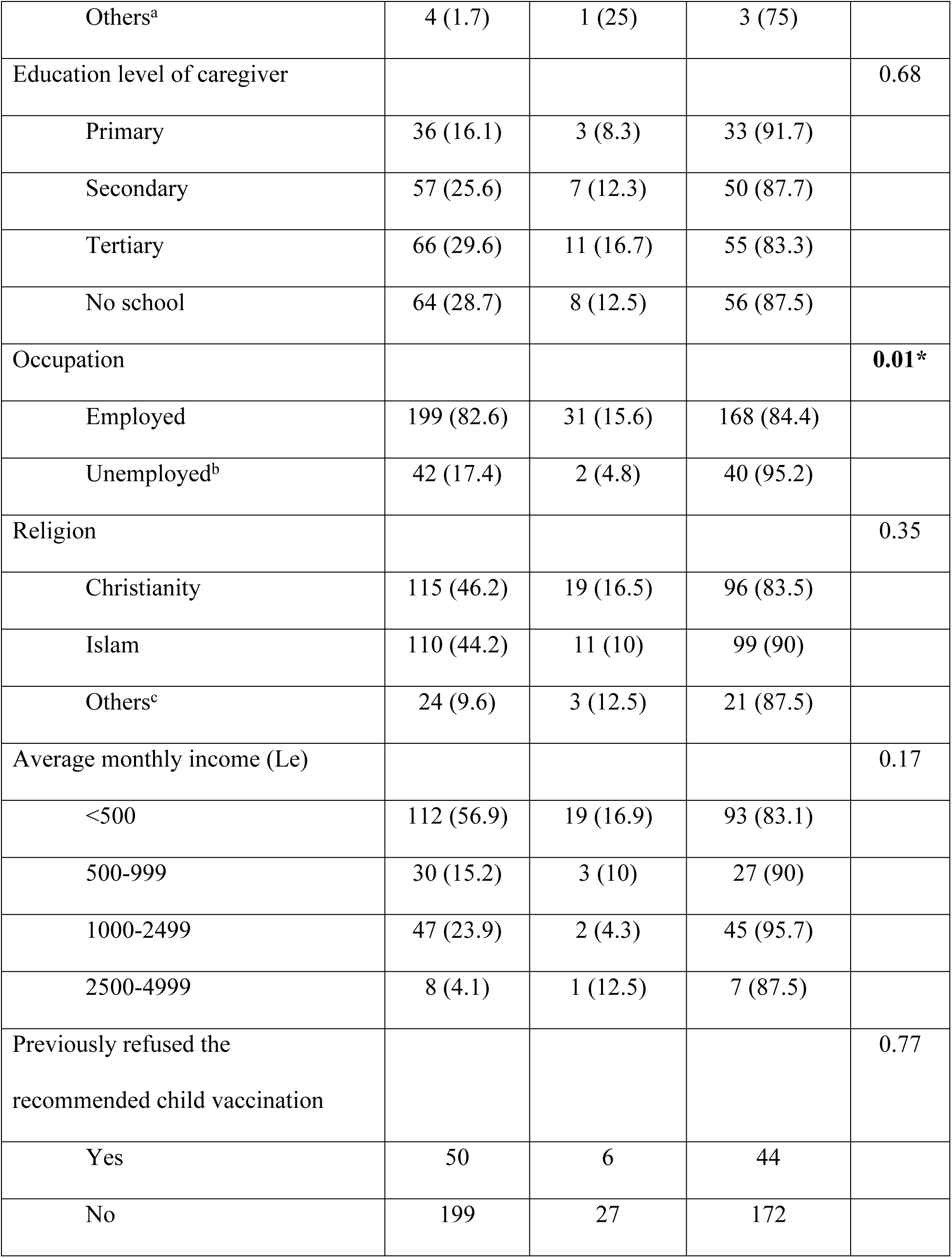
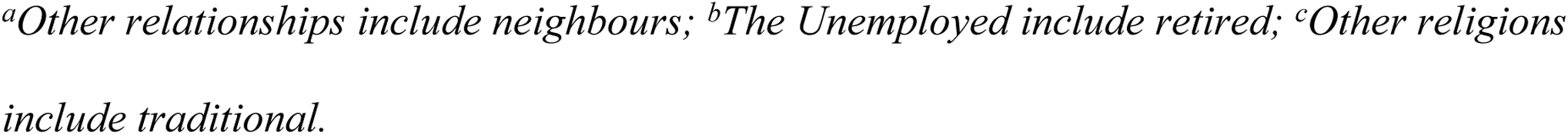
Baseline sociodemographic characteristics of 249 caregivers of adolescent girls living with HIV attending ART clinics in Sierra Leone by reported vaccination status.

| Variable | Overall<br>n = 249 | HPV vaccine status |  | p-value |
| --- | --- | --- | --- | --- |
|  |  | Vaccinated<br>n = 33 | Unvaccinated<br>n = 216 |  |
| Age of child, median (IQR) | 15 (12-17) | 16 (15-18) | 14 (11 -16) | **0.003\*** |
| Duration of HIV diagnosis, mean (SD) | 39.59 (46.5) | 17.79 (15.47) | 42.9 (48.74) | **0.004\*** |
| Age of caregiver |  |  |  | 0.22 |
| 18-30 | 60 (24.5) | 12 (20) | 48 (80) |  |
| 31-50 | 159 (64.9) | 20 (12.6) | 139 (87.4) |  |
| 51-70 | 20 (8.2) | 19 (95) | 1 (5) |  |
| 71-90 | 6 (2.4) | 0 | 6 (100) |  |
| Sex of caregiver |  |  |  | 0.32 |
| Male | 51 (20.6) | 9 (17.6) | 42 (82.4) |  |
| Female | 195 (78.6) | 42 (21.5) | 171 (87.7) |  |
| Relationship |  |  |  | **0.023\*** |
| Parent | 111 (47.8) | 16 (14.4) | 95 (85.6) |  |
| Sibling | 41 (17.7) | 3 (7.3) | 38 (92.7) |  |
| Extended family | 36 (15.5) | 9 (25) | 27 (75) |  |
| Grandparent | 29 (12.5) | 0 | 29 (100) |  |
| Family friend | 11 (4.7) | 2 (18.2) | 9 (81.8) |  |
| Others<sup>a</sup> | 4 (1.7) | 1 (25) | 3 (75) |  |
| Education level of caregiver |  |  |  | 0.68 |
| Primary | 36 (16.1) | 3 (8.3) | 33 (91.7) |  |
| Secondary | 57 (25.6) | 7 (12.3) | 50 (87.7) |  |
| Tertiary | 66 (29.6) | 11 (16.7) | 55 (83.3) |  |
| No school | 64 (28.7) | 8 (12.5) | 56 (87.5) |  |
| Occupation |  |  |  | **0.01\*** |
| Employed | 199 (82.6) | 31 (15.6) | 168 (84.4) |  |
| Unemployed<sup>b</sup> | 42 (17.4) | 2 (4.8) | 40 (95.2) |  |
| Religion |  |  |  | 0.35 |
| Christianity | 115 (46.2) | 19 (16.5) | 96 (83.5) |  |
| Islam | 110 (44.2) | 11 (10) | 99 (90) |  |
| Others<sup>c</sup> | 24 (9.6) | 3 (12.5) | 21 (87.5) |  |
| Average monthly income (Le) |  |  |  | 0.17 |
| <500 | 112 (56.9) | 19 (16.9) | 93 (83.1) |  |
| 500-999 | 30 (15.2) | 3 (10) | 27 (90) |  |
| 1000-2499 | 47 (23.9) | 2 (4.3) | 45 (95.7) |  |
| 2500-4999 | 8 (4.1) | 1 (12.5) | 7 (87.5) |  |
| Previously refused the recommended child vaccination |  |  |  | 0.77 |
| Yes | 50 | 6 | 44 |  |
| No | 199 | 27 | 172 |  |

<sup>a</sup>Other relationships include neighbours; <sup>b</sup>The Unemployed include retired; <sup>c</sup>Other religions include traditional.

### Overall Knowledge Score

The histogram (Figure 1) shows the distribution of participants’ knowledge scores as a percentage of correct responses, ranging from 0 to 10, and the percentage of participants for each knowledge score.

Overall, most participants have a low knowledge score, with a significant number having a score of 0. Most caregivers (40%) have a knowledge score of 0, and a smaller percentage (10%) have a knowledge score of 1. As the knowledge score increases, the percentage of participants decreases. Beyond the 4% correct mark, the percentage of participants drops significantly and remains low. The histogram shows a normal distribution of knowledge scores distributed around the median.

### Sources of Information and Knowledge of HPV Infection, Cervical Cancer and Its Prevention

The highest percentage among all the information sources comes from HIV service providers (39.4%). Community members and religious leaders follow this, each accounting for approximately 21.2% of information sources (Figure 2).

When HPV vaccine knowledge scores were assessed across various sources of information among caregivers of AGLHIV, a statistically significant (p = 0.039) difference in knowledge scores was observed across information sources. Healthcare Workers were associated with the highest knowledge scores, while community members were linked to the poorest knowledge (Table 3).

**Table 3.** Kruskal-Wallis Test Results for Differences in Knowledge Scores by HPV Vaccine Information Source.

| Information Source | *n* | Rank Sum |
| --- | --- | --- |
| Healthcare workers | 26 | 889.50 |
| Family member | 4 | 176.00 |
| Social media | 6 | 185.00 |
| Mass media | 13 | 529.50 |
| Community member | 1 | 16.50 |
| Religious leaders | 14 | 298.50 |
| Test Statistic:<br>$\chi^2(1) = 12.87, p=0.045$(unadjusted for ties)<br>$\chi^2(5) = 13.29, p=0.039$(adjusted for ties) |  |  |

Table 4 presents the proportion of responses to selected questions regarding knowledge of the HPV infection, vaccine and cervical cancer across vaccination status. Vaccinated caregivers consistently demonstrated better knowledge across most domains, though overall awareness remained low. Only 32.1% (80/249) of caregivers had heard of HPV infection. Vaccinated caregivers were significantly more likely to report awareness (40%, 32/80) than unvaccinated caregivers (60%, 48/80). Notably, 99.4% (168/169) of caregivers who had not heard of HPV were unvaccinated. Less than half (41.1%, 102/249) knew about cervical cancer, with vaccinated caregivers marginally more informed (28.4%, 29/102) than unvaccinated caregivers (71.6%, 73/102).

**Table 4.** Reproductive health knowledge on HPV and cervical cancer among 249 caregivers of adolescents living with HIV, stratified by vaccination status.

|  | Total<br>n (%) | Vaccinated<br>n (%) | Unvaccinated<br>n (%) |
| --- | --- | --- | --- |
| Heard about HPV infection |  |  |  |
| Yes | 80 (32.1) | 32 (40) | 48 (60) |
| No | 169 (67.9) | 1 (0.6) | 168 (99.4) |
| Heard about cervical cancer |  |  |  |
| Yes | 102 (41.1) | 29 (28.4) | 73 (71.6) |
| No | 146 (58.9) | 4 (2.7) | 142 (97.3) |
| Cervical cancer is preventable |  |  |  |
| Yes | 79 (31.9) | 29 (36.7) | 50 (63.3) |
| No | 9 (3.6) | 0 | 9 (100) |
| I don't know | 160 (64.5) | 4 (2.5) | 156 (97.5) |
| Early sexual debut increases the risk of cervical cancer |  |  |  |
| Yes | 71 (28.7) | 25 (35.2) | 46 (64.8) |
| No | 13 (5.3) | 3 (23) | 10 (76.9) |
| I don't know | 163 (66) | 5 (3.1) | 158 (96.9) |
| HPV infection causes cervical cancer |  |  |  |
| Yes | 60 (24.4) | 31 (51.7) | 29 (48.3) |
| No | 3 (0.12) | 1 (33.3) | 2 (66.7) |
| I don't know | 183 (74.4) | 48 (26.2) | 135 (73.8) |
| Oral contraceptive increases the risk of cervical cancer |  |  |  |
| Yes | 36 (16.3) | 17 (47.2) | 19 (52.8) |
| No | 40 (18.1) | 11 (27.5) | 29 (72.5) |
| I don't know | 145 (65.6) | 4 (2.8) | 141 (97.2) |
| Multiple sexual partners increase risk of cervical cancer |  |  |  |
| Yes | 72 (30) | 23 (31.9) | 49 (68.1) |
| No | 19 (7.9) | 9 (47.4) | 10 (52.6) |
| I don't know | 149 (62.1) | 1 (0.7) | 148 (99.3) |
| Smoking increases the risk of cervical cancer |  |  |  |
| Yes | 48 (19.6) | 17 (35.4) | 31 (64.6) |
| No | 41 (16.7) | 12 (29.3) | 29 (70.7) |
| I don't know | 156 (63.7) | 4 (2.6) | 152 (97.4) |
| HIV increase the risk of cervical cancer |  |  |  |
| Yes | 85 (35.4) | 28 (32.9) | 57 (67.1) |
| No | 31 (12.9) | 1 (3.2) | 30 (96.8) |
| I don't know | 124 (51.7) | 3 (2.4) | 121(97.6) |

Only 31.9% (79/249) correctly identified cervical cancer as preventable. Vaccinated caregivers demonstrated slightly higher awareness (36.7%, 29/79 vs. 63.3% unvaccinated). A minority (24.4%, 60/249) knew HPV causes cervical cancer. Vaccinated caregivers were twice as likely to recognize this link (51.7%, 31/60 vs. 48.3% unvaccinated). However, 74.4% (183/249) reported uncertainty.

Early sexual debut was correctly identified as a risk factor (28.7%), with vaccinated caregivers more informed (35.2%, 25/71 vs. 64.8% unvaccinated). Similarly, 30% (72/249) acknowledged that having multiple sexual partners is a risk, with vaccinated caregivers again more aware (31.9%, 23/72 vs. 68.1% unvaccinated). Only 35.4% (85/249) knew HIV increases cervical cancer risk, but only 32.9% (28/85) of these were vaccinated.

With regards to misconceptions and knowledge, oral contraception (16.3%) was incorrectly believed to increase cervical cancer risk. While 19.6% (48/245) recognized smoking as a risk factor, over 60% of caregivers reported uncertainty across questions about risk factors highlighting pervasive knowledge gaps.

### Reason for HPV non-vaccination

The reasons for HPV non-vaccination and unwillingness toward future vaccination among 233 caregivers of AGLHIV in Sierra Leone are presented in Table 5. Over two-thirds (66.1%, 154/233) of non-vaccination cases stemmed from caregivers being unaware of the HPV vaccine. Religious beliefs (25%, 18/72) and fear (12.5%, 9/72) dominated future unwillingness while competing daily priorities and missed vaccination opportunities indicate systemic issues in healthcare access and program implementation.

**Table 5.** Reasons for HPV non-vaccination and unwillingness towards future vaccination among caregivers of AGLHIV in Sierra Leone.

| Reason for HPV non-vaccination, n = 233 | n (%) |
| --- | --- |
| Unaware | 154 (66.1) |
| Concerns about side effects | 11 (4.7) |
| Not Visited | 24 (10.3) |
| Absent on the day of school vaccination | 15 (6.4) |
| Underage or Overage | 4 (1.7) |
| Non-consent spouse | 1 (0.4) |
| Daily activities (stress) | 24 (10.3) |
| Reason for unwillingness to take HPV vaccination in the future, n = 72 |  |
| Fear | 9 (12.5) |
| Travel | 1 (1.4) |
| Beliefs (e.g, Vaccines are ineffective, makes me uncomfortable) | 44 (61.1) |
| My religion does not support vaccination | 18 (25) |

### Factors Associated with HPV Vaccination

The association between caregivers’ education level and both HPV vaccine knowledge and willingness among caregivers of adolescents living with HIV in Sierra Leone is presented in Tables 6 and 7.

**Table 6.** Knowledge Score, Willingness Distribution and Association with Education Level.

| Variable |  | p-value (x2) |
| --- | --- | --- |
| **Education Level** | **Adequate knowledge** |  |
|  | **n (%)** |  |
| No school | 8/64 (12.5) |  |
| Primary | 3/36 (8.3) |  |
| Secondary | 8/57 (14) |  |
| Tertiary | 30/66 (45.5) | **<0.001\* (30.58)** |
| **Education Level** | **Willingness** | 0.295 |
|  | **n (%)** |  |
| No school | 22/64 (34.4) |  |
| Primary | 17/36 (47.2) |  |
| Secondary | 23/57 (40.4) |  |
| Tertiary | 33/66 (50) |  |

**Table 7.** Factors associated with the attitude of HPV and cervical cancer among caregivers of adolescents living with HIV in Sierra Leone.

| Variable | Willingness towards HPV vaccine |  | p-value | COR (95% CI) | p-value |
| --- | --- | --- | --- | --- | --- |
|  | Yes (n, %) | No, (n, %) |  |  |  |
| Age of caregiver |  |  | 0.059 | 0.293 (0.064-1.338) | 0.113 |
| 18-30 | 28 (46.7) | 32 (53.3) |  |  |  |
| 31-50 | 60 (37.7) | 99 (62.3) |  |  |  |
| 51-70 | 13 (65) | 7 (35) |  |  |  |
| 71-90 | 4 (66.7) | 2 (33.3) |  |  |  |
| Sex of caregiver |  |  | 0.075 | 3.138 (0.522-18.856) | 0.211 |
| Male | 28 (54.9) | 23 (45.1) |  |  |  |
| Female | 78 (40) | 117 (60) |  |  |  |
| Relationship |  |  | **0.001\*** | 0.428 (0.199-0.916) | **0.029\*** |
| Grandparent | 9 (31) | 20 (69) |  |  |  |
| Sibling | 22 (53.7) | 19 (46.3) |  |  |  |
| Extended family | 25 (69.4) | 11 (30.6) |  |  |  |
| Parent | 37 (33.3) | 74 (66.7) |  |  |  |
| Family friend | 1 (25) | 3 (75) |  |  |  |
| Education level of caregiver |  |  | 0.298 | 1.185 (0.437-3.217) | 0.739 |
| Primary | 17 (47.2) | 19 (52.8) |  |  |  |
| Secondary | 23 (40.4) | 34 (59.6) |  |  |  |
| Tertiary | 33 (50) | 33 (50) |  |  |  |
| No school | 42 (65.6) | 22 (34.4) |  |  |  |
| Occupation |  |  | 0.341 | 0.408 (0.128-1.301) | 0.130 |
| Employed | 78 (42.6) | 105 (57.4) |  |  |  |
| Unemployed<sup>b</sup> | 22 (40.7) | 32 (59.3) |  |  |  |
| Religion |  |  | 0.852 | 1.778 (0.462-1.683) | 0.403 |
| Christianity | 47 (40.9) | 68 (59.1) |  |  |  |
| Islam | 49 (44.5) | 61 (55.5) |  |  |  |
| Others<sup>c</sup> | 10 (41.7) | 14 (58.3) |  |  |  |
| Average monthly income (Le) |  |  | 0.129 | 1.692 (0.551-1.520) | 0.358 |
| <500 | 53 (47.3) | 59 (52.7) |  |  |  |
| 500-999 | 7 (23.3) | 23 (76.7) |  |  |  |
| 1000-2499 | 20 (42.6) | 27 (57.4) |  |  |  |
| 2500-4999 | 3 (37.5) | 5 (62.5) |  |  |  |
| Information source |  |  |  | 0.732 (0.383-1.401) | 0.347 |

A significant association was observed between education level and adequate HPV vaccine knowledge (p = 0.001). Caregivers with tertiary education demonstrated the highest proportion of adequate knowledge (45.5%, 30/66), far exceeding other groups (Table 6).

The caregiver’s relationship with AGLHIV was observed to show a strong association with willingness to vaccinate (p = 0.029). Extended family members exhibited the highest willingness (69.4%, 25/36), followed by Siblings (53.7%, 22/41). Male caregivers reported higher willingness (54.9%, 28/51) than female caregivers (40%, 78/195), but this difference was not significant (p=0.211; aOR = 3.138, 95% CI: 0.522–18.856).

## Discussion

In this study of 249 caregivers of adolescent girls living with HIV attending HIV clinics in tertiary hospitals in Sierra Leone, we assessed knowledge, attitudes and willingness to acceptHPV vaccine for AGLHIV.

This study highlights four key findings. First, caregivers showed substantial gaps in knowledge about HPV, cervical cancer, and vaccination. Second, lack of awareness about vaccination campaigns and service locations was a major barrier to uptake. Third, healthcare workers played a crucial role in information dissemination, with caregivers informed by them demonstrating greater awareness. Lastly, the caregiver’s relationship to the AGLHIV was significantly associated with willingness to vaccinate (p = 0.029) .Consistent with studies in low-resource settings, caregivers demonstrated significant gaps in HPV and cervical cancer knowledge [20]. Only 24.4% recognized HPV as a cause of cervical cancer, and 64.5% were unaware of its preventability. These rates are lower than those reported in Kenya (35% awareness) [21] and Nigeria (28%) [22–24], likely due to Sierra Leone’s limited cancer prevention programs. Misconceptions, such as linking oral contraceptives to cervical cancer (16.3%), echo findings in many parts of SSA, where cultural beliefs often conflate contraception with disease risk [25–27]. The pervasive “I don’t know” responses (60–74%) underscore an urgent need for culturally tailored education campaigns, particularly targeting unvaccinated caregivers who constituted 99% of this uncertain cohort.

While tertiary-educated caregivers exhibited the highest knowledge, their willingness to vaccinate was only marginally higher than less-educated groups. This paradox mirrors findings in SSA, where educated caregivers’ vaccine hesitancy stemmed from distrust in government-led programs [28–30]. Similarly, extended family members—despite limited formal education—showed the highest willingness, likely due to their closer engagement with adolescents’ healthcare needs compared to grandparents. These results align with the Health Belief Model, where perceived benefits and cues to action (e.g., familial responsibility) often outweigh knowledge alone [31–33].

Unawareness was the primary barrier to vaccination, reflecting systemic gaps in Sierra Leone’s HPV vaccine rollout, which relies heavily on school-based programs inaccessible to out-of-school AGLHIV [34]. Logistical challenges, such as missed vaccination days, parallel barriers identified in Malawi, where mobility constraints hindered clinic access [35]. Vaccine attitudes in Sierra Leone are closely influenced by religious beliefs. Studies across Sierra Leone, Liberia, and Guinea reported lower Hepatitis B vaccine completion among children of Muslim mothers compared to Christians. Other findings also show that children with Muslim caregivers are more likely to miss routine vaccinations. While these patterns are not universal among Muslims, hesitancy may also stem from broader issues such as inter-religious tensions or the social exclusion of minorities, which can deepen mistrust in health systems. It is important, however, not to generalize vaccine rejection within Muslim communities[36, 37]. Addressing these requires decentralizing vaccination services and collaborating with religious institutions, as demonstrated in Rwanda’s HPV vaccine scale-up [38].

Healthcare workers emerged as pivotal information sources, with caregivers relying on them demonstrating higher knowledge. This aligns with WHO recommendations prioritizing healthcare worker training in LMICs [39]. However, most caregivers (41.2%) receive information through the mass media (radio and television), indicating that caregivers’ knowledge levels varied significantly depending on their primary source of HPV vaccine information. The limited impact of mass media and social media contrasts with settings where digital platforms improved vaccine literacy [40–42], suggesting Sierra Leone’s infrastructural limitations (e.g., low internet penetration) require alternative strategies, such as radio-based campaigns.

To improve HPV vaccine uptake among AGLHIV in Sierra Leone, multiple approaches rooted in community trust and systemic integration is critical [43–46]. First, empowering extended family members and healthcare workers as peer educators can strengthen messaging through established social networks, ensuring information reaches caregivers in familiar, trusted settings. To combat pervasive myths, locally tailored radio dramas and town hall discussions led by healthcare professionals can emphasize vaccine safety and efficacy while directly addressing community concerns. Finally, integrating HPV vaccination into HIV care protocols would streamline service delivery, leveraging existing infrastructure and trust in HIV programs to bolster vaccine acceptance.

This study has several limitations that warrant consideration. First, the overrepresentation of tertiary-educated caregivers and urban clinics in the sample may restrict the generalizability of findings to rural populations and caregivers with lower educational attainment. Second, the cross-sectional design precludes causal inference; while associations between variables were identified, longitudinal studies are needed to establish temporal relationships and assess how knowledge translates into vaccine uptake over time. Third, unmeasured confounders, such as healthcare access, community stigma, and socioeconomic dynamics, were not evaluated but may critically shape caregivers’ willingness to vaccinate.

## Conclusion

Improving HPV vaccine uptake among AGLHIV in Sierra Leone demands a dual focus on education and systemic equity. Concerns about vaccination are frequently dismissed as mere misconceptions. However, acknowledging these fears as valid and engaging with them directly is essential for building trust in public health systems. Traditional media and in-person engagement are key in shaping how individuals interpret online information, helping them distinguish between accurate content and misinformation. By addressing knowledge gaps, leveraging trusted community actors, and dismantling structural barriers, Sierra Leone can mitigate cervical cancer disparities in this vulnerable population.

## List of Abbreviations

AGLHIV: Adolescent girls living with Human Immunodeficiency Virus
ART: Antiretroviral Therapy
HIV: Human Immunodeficiency Virus
HPV: Human papillomavirus
PACV: Parents’ Attitude about Childhood Vaccination
STI: Sexually transmitted infection
WHO: World Health Organization

## Acknowledgments

We thank all ICHAD members at Brown School at Washington University in St. Louis, the program managers for the ACHIEVE program, Laura Peer, Chelsea Hand-Sheridan, Bethel Mandefro, Rebecca Esliker, the University of Makeni, the Caregivers, the ART clinics staff, and the Fogarty International Centre.

## Author Contributions

DFJ and JD conceptualized and designed the research study. DFJ, MNK, UB, WOA, DS, RS, MMB, DS, PMDJ, collected the required data. DFJ and MB conducted data analysis, interpreted the results and drafted the initial version of the manuscript. All authors have reviewed multiple drafts, provided significant edits and feedback, and approved the final version for submission. JD, as the ACHIEVE program mentor, oversaw the entire project, providing a critical review, guidance, and leadership throughout the research process.

## Data availability statement

Data will be made available upon request to the corresponding author.

## Supplementary

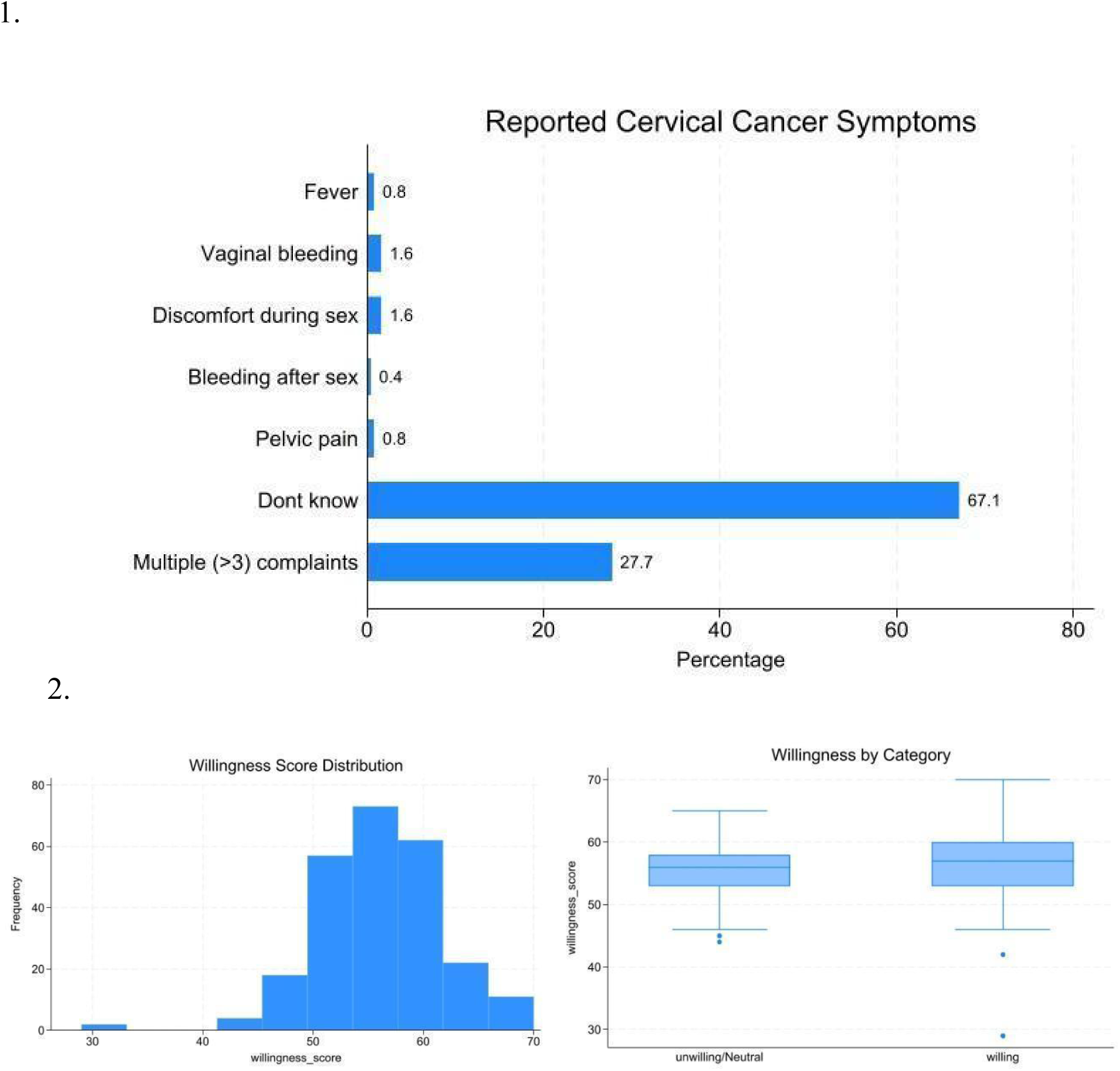

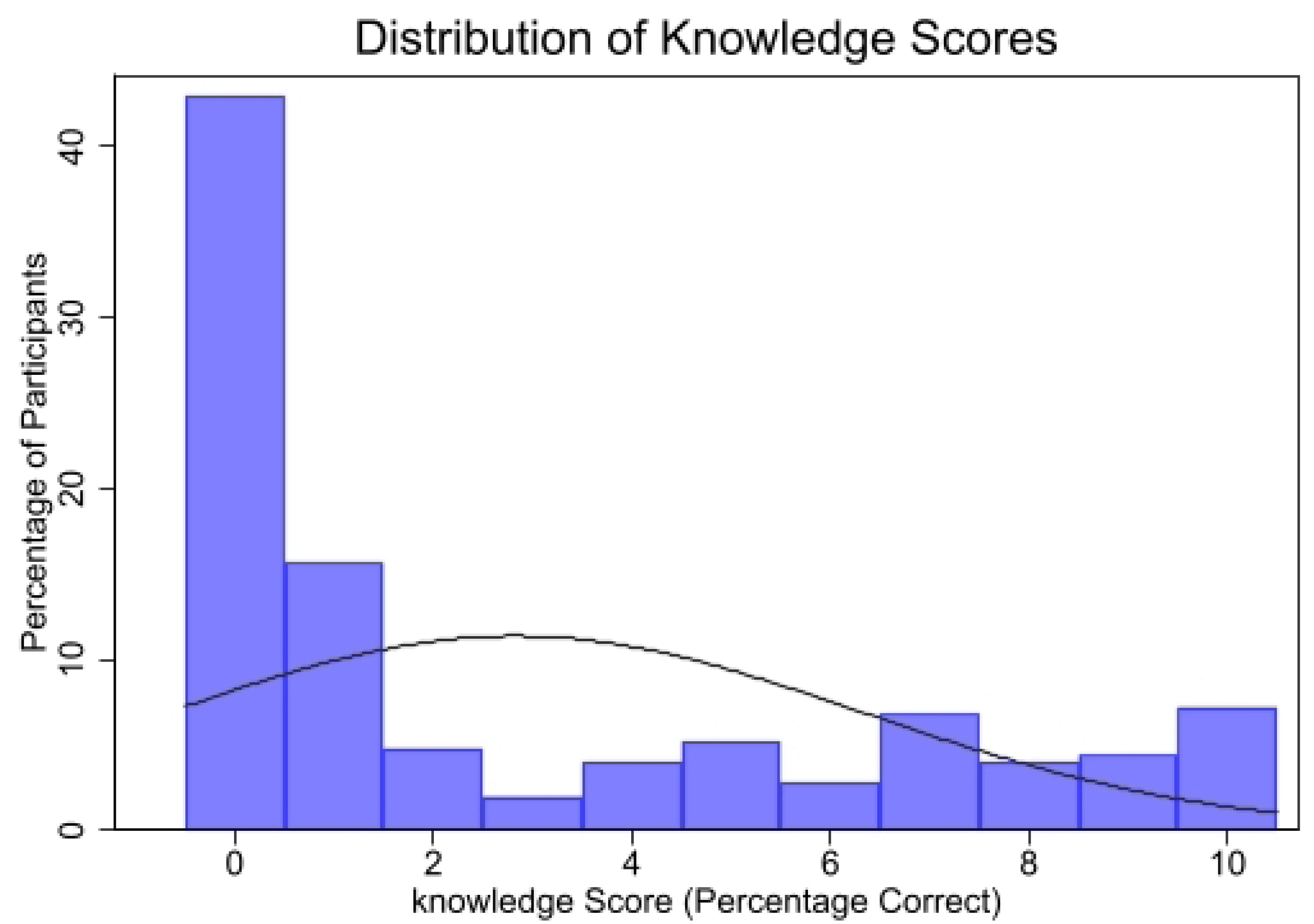

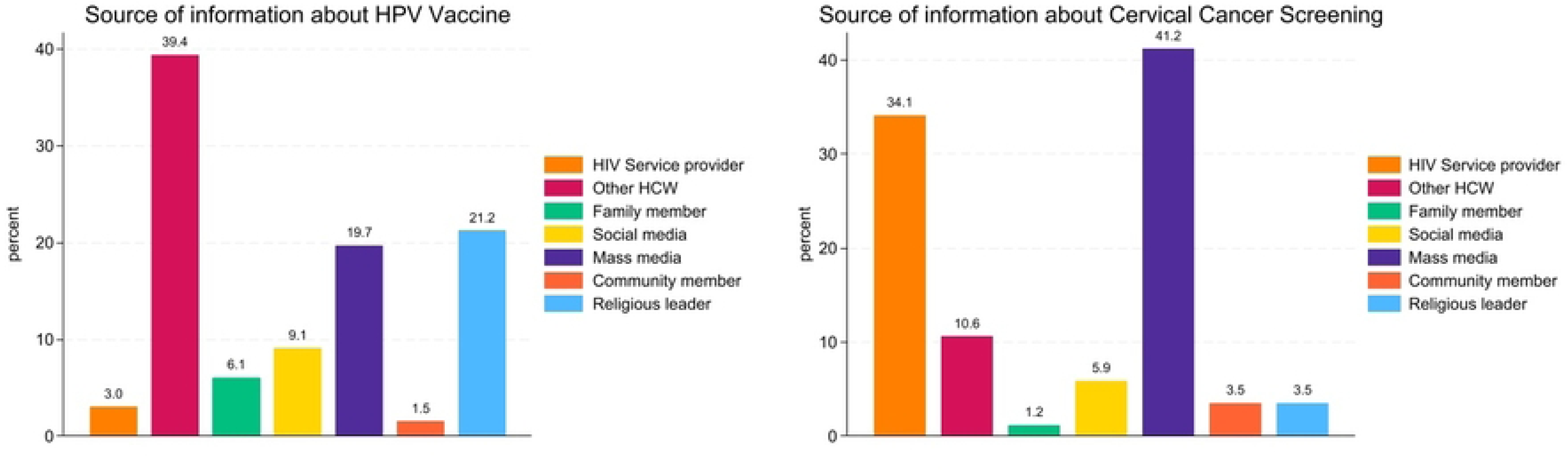

